# An Integrated Neural Network and Evolutionary Algorithm Approach for Liver Fibrosis Staging: Can Artificial Intelligence Reduce Patient Costs?

**DOI:** 10.1101/2024.03.05.24303786

**Authors:** Ali Nazarizadeh, Touraj Banirostam, Taraneh Biglari, Mohammadreza Kalantarhormozi, Fatemeh Chichagi, Amir Hossein Behnoush, Mohammad Amin Habibi, Ramin Shahidi

**Author notes:** **Corresponding author’s contact information:** Ramin Shahidi, MD. School of Medicine, Bushehr University of Medical Sciences, Bushehr, Iran, Address: Bushehr University of Medical Sciences, Moallem St, Bushehr, Iran. Tell. **Data availability statement:** The data is available within the article or is achievable through the request of the corresponding author. **Funding statement:** The present study has not received any funding. **Ethics approval statement:** This article contains no patient-identifying information, so ethical approval is not required.

## Abstract

**Background:** Liver fibrosis is important in terms of staging, and liver biopsy is the gold standard diagnostic tool. We aim to design and evaluate an artificial neural network (ANN) method by taking advantage of the Teaching Learning Based Optimization (TLBO) algorithm for the prediction of liver fibrosis stage in blood donors and hepatitis C.

**Method:** We proposed a method based on a selection of machine learning classification methods including Multi Layers Perceptron neural network (MLP), Naive Bayesian (NB), decision tree, and deep learning. Initially, the Synthetic minority oversampling technique (SMOTE) was performed to address the imbalance of the dataset. Afterward, the integration of MLP and TLBO was implemented.

**Result:** We proposed a novel algorithm that reduced the number of required patient features to 7 inputs. The accuracy of MLP using 12 features is 0.903, while the accuracy of the proposed MLP with the TLBO method is 0.891. Besides, the diagnostic accuracy in all methods, except the model designed with the Bayesian Network, increased when the SMOTE balancer was applied.

**Conclusion:** The Decision tree deep learning methods showed the highest levels of accuracy with 12 features. Interestingly, with the use of TLBO and 7 features, the MLP reached a 0.891 accuracy rate which is quite satisfying compared with similar studies. The proposed model provided great diagnostic accuracy by reducing the required properties from the samples without reducing the accuracy. The results of our study showed that the recruited algorithm of our study was more straightforward, with lower required properties and similar accuracy.

## Introduction

Chronic liver diseases are among the top causes of death worldwide, accounting for two million deaths worldwide annually, and are responsible for about 3.5% of total deaths. Half of this number can be attributed to cirrhosis complications and remaining to the viral hepatitis and hepatocellular carcinoma (1). Among them, the hepatitis C virus (HCV), considered to be the main cause of liver cancer, is a blood-borne virus spreading via unsafe drug injection, blood transfusion, and unsafe sexual practices (2). While 95% of patients are manageable, it can lead to liver cirrhosis or cancer. After six months, the chronic phase of the disease starts and influences the liver and causes inflammatory mediators production which makes the liver produce fibrous proteins to repair the damage (2). This is called liver fibrosis which results in less blood flow to the liver cells and eventually leads to necrosis (3).

The METAVIR score has been designed to stage the fibrosis in chronic hepatitis C, with 0 representing no fibrosis and 4 showing severe cirrhosis or scarring (4). However, this scoring system is based on liver biopsy, the gold standard for fibrosis staging. However, it has some drawbacks which may limit its use. First, it is an invasive method for diagnosis which can lead to several complications, including biliary peritonitis, hemorrhage, and even pneumothorax (5). In addition, sampling errors and variations in histologic evaluations are inevitable (6). Finally, a biopsy cannot be utilized as a usual assessment tool for prognosis and follow-ups. Serum biomarkers and imaging modalities are some of the alternatives for liver biopsy (7). However, most of these options are associated with a lack of high sensitivity and specificity, such as the aspartate aminotransferase-to-platelet ratio index (APRI) with 69% sensitivity and 77% specificity (8).

The applications of Artificial Intelligence (AI) in medicine have grown rapidly in recent years (9). It can be used for prediction by classification methods using data mining and machine learning (ML) algorithms. Several AI projects have been conducted in order to predict staging of hepatitis C, including some ML methods (10-12). However, most of these have used a small number of parameters or could not achieve high accuracy.

Artificial neural network (ANN) is one of the most commonly used ML algorithms which mimic the human brain in processing inputs and converting them to an output via some hidden layers (13). The Teahing Learning Based Optimization algorithm (TLBO) is a new population-based optimization algorithm which was proposed in 2011 and can find optimal solutions in a shorter period of time (14, 15). It is inspired by a number of learners taught by a teacher. Each of the learners is considered a solution with its dimensions representing the parameters of the objective function of the given optimization problem (16). Herein, we aim to prepare and investigate a Machine learning algorithm on a pre-set dataset by utilizing ANN with TLBO optimization for feature selection to predict the staging of liver fibrosis in blood donors and hepatitis C patients.

## Materials and method

### Data mining and data source

Data mining is extracting data to analyze it more accurately and efficiently to improve decision-making efficiency. For our analysis, a dataset of blood donors and HCV patients obtained from the UCI machine learning repository (17) is used for our analysis.

### Study Outcome

The main outcome of the study was liver fibrosis staging, for which we compared different ML-based prediction models. In a random assignment process, the data was divided into train and test subsets (70% and 30%, respectively). Evaluation of the models was based on test data.

### Data Balancing

Due to the imbalanced data regarding positive and negative HCV subjects, we used the Synthetic minority oversampling technique (SMOTE). It identifies the minority group’s k-nearest neighbors, then selects a set of neighbors which generates new data (18). Moreover, the min-max method and substituting missing data with the mean were used to balance the dataset.

### Teaching Learning Based Optimization algorithm (TLBO)

We used TLBO (19) to lower the number of features. This algorithm comprises two phases: 1) the Teacher phase, in which the learners get the knowledge from the teacher as the best solution available, and 2) the Learner phase, in which learners learn through interacting with each other (20, 21). This method is based on a teacher’s influence on students’ output in a class, which means it works based on the man. The teacher is the best way of selecting, and students’ output is defined according to the mean of each selection method. In other words, this method compares the means of every random selection technique and chooses the highest one. The most significant advantage of this method is finding the optimal solution in a short computational time (14).

### Machine learning algorithms

After data preparation, four algorithms were implemented in the balanced dataset: Multi Layers Perceptron neural network (MLP), Naive Bayesian, decision tree and deep learning. Afterward, the models were compared. Our proposed method was the integration of TLBO and MLP. All algorithms were implemented in the Keras environment.

### Multi Layers Perceptron neural network (MLP)

MLP is an extended neural network in which neurons are strongly connected. This study contains an input layer containing 12 neurons, an output layer with five neurons, and one hidden layer. We applied the function Ruel (f(x)= max (x,0)) to enhance the network’s learning pace. Furthermore, layer dropout was applied in the middle layer. In every election, only neurons with a higher level of possibility (p-value >0.05) were used, preventing overfitting. In this study, the MLP technique used 70% of the dataset cases as training data and 30% as testing.

### Naive Bayesian

This classification algorithm is based on the Bayes theorem with the objectivity assumption between the predictors. In this method, the dataset is considered as input on which the analysis is conducted. Afterward, the class label will be predicted using Bayes’ Theorem, and the probability of class in input data is calculated, which can help predict the class for the unknown data sample. This classification method can be suitable, especially for large datasets.

### Deep Learning

Deep learning is a subset of machine learning techniques that focuses on training artificial neural networks with multiple layers, or “deep” networks. Unlike traditional machine learning models, deep learning models can automatically learn patterns and features from the data, making them suitable for complex tasks such as image recognition and natural language processing. In our research, deep learning was employed as one of the machine learning algorithms to enhance the accuracy of liver fibrosis staging predictions.

### Decision tree

This is one of the most commonly used techniques for classifying algorithms. This algorithm’s basis is dividing data into subcategories based on a series of questions. The starting point is the root node, which is the tree’s root centered on the highest entropy and contains all samples. Each node is then split into secondary or leaf nodes in binary form or a multi-split. This model is like a tree structure that includes a group of nodes. It contains all decision nodes (splits node with the condition) and leaf nodes. The workflow of the dataset processing is illustrated in Figure 1.

**Figure 1.**
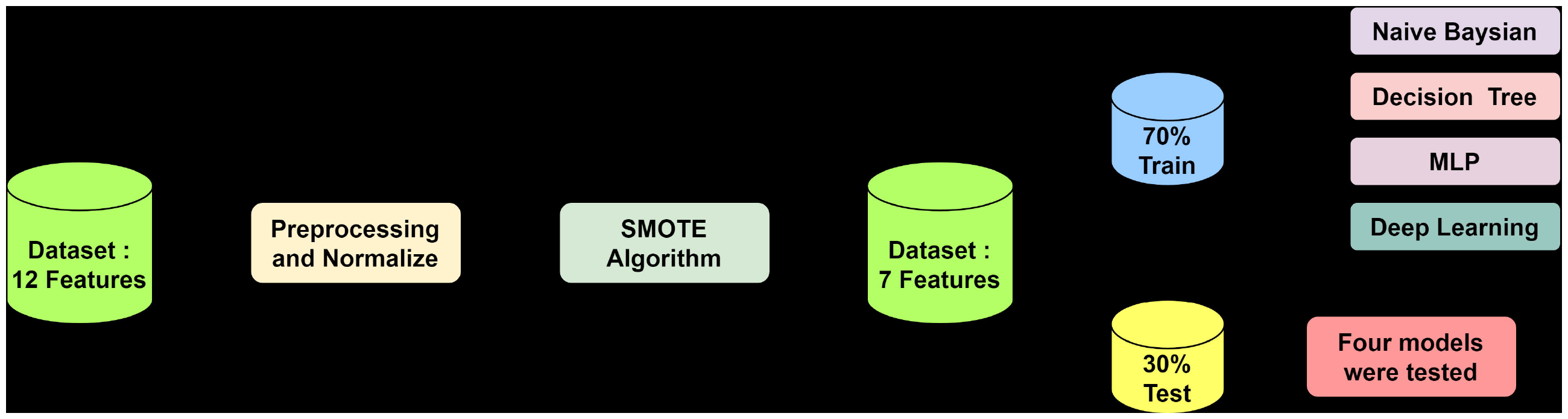

### Algorithm Evaluation

The primary metric for evaluating the models was accuracy, defined as the number of correct predictions by the total number of samples. In addition to accuracy, we also reported three essential metrics for a comprehensive evaluation of the model’s performance:

#### Precision

This metric helps us understand the proportion of true positive predictions (correctly identified liver fibrosis cases) out of all the positive predictions made by the model.

#### Recall

Also known as sensitivity or true positive rate, recall gauges the model’s capacity to identify all actual positive cases. In the context of liver fibrosis, recall measures how effectively the model detects individuals with the condition, reducing the chances of false negatives.

#### F1 Score

The F1 score is a balanced measure that considers both precision and recall, providing a single value that reflects the model’s overall performance. It’s particularly useful when there’s an imbalance between positive and negative cases in the dataset. A high F1 score suggests a model that effectively identifies cases while minimizing false positives and false negatives.

## Results

Our study focused on developing and evaluating machine learning-based models for predicting liver fibrosis stages in blood donors and hepatitis C patients. We employed a dataset from the UCI Machine Learning Repository, which included 615 records with an average age of 47.29 ±9.993 years, all of which had the diagnosis of serology or histopathology negative or positive HCV; 75 patients with positive HCV test (including 24 Hepatitis, 21 Fibrosis, and 30 Cirrhosis), each characterized by various clinical and demographic features. We removed 7 cases due to suspicious diagnosis. Twelve data features were recruited for this study: age, sex, albumin blood test (ALB), alkaline phosphatase (ALP), alanine transaminase (ALT), aspartate transaminase (AST), acetylcholinesterase (CHE), cholesterol (CHOL), creatinine (Cr), bilirubin (BIL), Gamma-Glutamyl Transferase (GGT), and proteins. The primary objective was to identify an optimized subset of features and assess the performance of machine learning algorithms in accurately predicting liver fibrosis stages. Table 1 provides a summary of patients’ characteristics in our dataset.

**Table 1:** Summary of Patients’ Characteristics.

| Features | Range | Mean ± SD | Missing |
| --- | --- | --- | --- |
| Age | 19-77 | 47.29 ± 9.993 | 0 |
| Sex | -- | 237 Female and 371 Male | 0 |
| ALP | 11.3-416.6 | 67.821 ± 25.2744 | 18 |
| ALT | 0.9-258 | 27.601 ± 21.2275 | 1 |
| AST | 12-324 | 34.369 ± 32.6224 | 0 |
| ALB | 20-82.2 | 41.819 ± 5.4067 | 1 |
| CHOL | 1.43-9.67 | 5.3788 ± 1.1194 | 10 |
| Cr | 8-1079 | 81.51 ± 49.721 | 0 |
| BIL | 1.8-25.4 | 11.474 ± 19.7706 | 0 |
| GGT | 4.5-650.9 | 38.244 ± 51.9532 | 0 |
| CHE | 1.42-16.41 | 8.2049 ± 2.1684 | 0 |
| Proteins | 51-90 | 72.253 ± 4.9323 | 1 |

### Feature Selection and Algorithm Performance

Our primary focus was to assess the performance of different machine learning algorithms in predicting liver fibrosis. We implemented the following algorithms: Multi Layers Perceptron neural network (MLP), Naive Bayesian, decision tree and deep learning. The models were compared based on their accuracy in predicting liver fibrosis stages.

In our study, we initially employed the Synthetic Minority Oversampling Technique (SMOTE) to address the challenge of imbalanced data regarding positive and negative HCV subjects. SMOTE was applied to create synthetic instances in the dataset. The results of employing the SMOTE method are shown in Table 2. The introduction of SMOTE led to an improvement in diagnostic accuracy across all methods. However, the diagnostic accuracy of the Bayesian Network model decreased after applying SMOTE, which was consistent with findings from previous studies using the same dataset. The highest accuracy was observed in the deep learning and decision tree, while the Bayesian Network model exhibited the lowest accuracy.

**Table 2:** Comparing the Diagnostic Accuracy of models with and without Using SMOTE.

| Algorithm | Accuracy without SMOTE | Accuracy with SMOTE |
| --- | --- | --- |
| Naive Bayesian | 0.870 | 0.855 |
| Decision Tree | 0.897 | 0.962 |
| MLP | 0.902 | 0.956 |
| Deep Learning | 0.908 | 0.966 |

Subsequently, we used the Teaching Learning Based Optimization (TLBO) algorithm to reduce the number of features from the initial set of 12. The selected features for our predictive models included Age, Albumin Blood Test (ALB), Alkaline Phosphatase (ALP), Alanine Transaminase (ALT), Aspartate Transaminase (AST), Cholesterol (CHOL), and Proteins.

The diagnostic accuracy of MLP algorithms trained with different features is provided in Table 3. The MLP model using all 12 features achieved an accuracy of 0.903, while the MLP model with TLBO-selected seven features maintained a high accuracy of 0.891. Notably, the reduction in features did not significantly compromise accuracy, indicating the effectiveness of feature selection in optimizing predictive performance. Additionally, the authors chose four blood indices at random to evaluate the power of random blood indices on liver fibrosis staging. Blood characteristics chosen at random included Gamma-Glutamyl Transferase (GGT), proteins, creatinine (Cr), and cholesterol (CHOL). However, it is important to note that this method did not achieve a high level of accuracy in predicting the liver fibrosis stage.

**Table 3:**
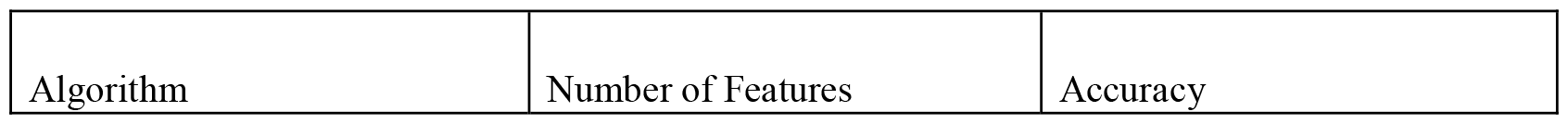
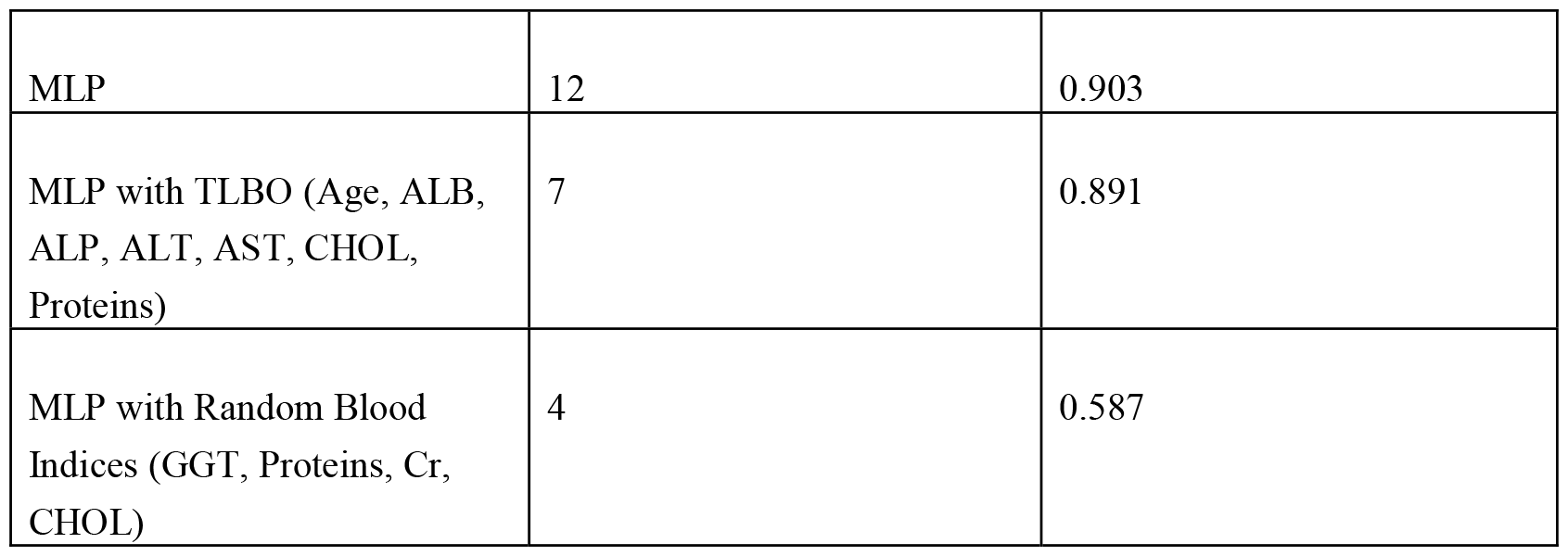
The Diagnostic Accuracy of MLP with different features.

Additionally, there are other useful algorithms such as Support Vector Machine (SVM) and random forests which are not reported in this paper due to less accuracy or precision in our initial analysis of this dataset and due to space limitations and the specific objective of our study, we just focused on the most accurate algorithms.

## Discussion

Given the high importance of data mining in medical science, numerous research has been done in this field. In earlier research on the subject of liver complications, the integration of the TLBO’s innovative algorithm has not been accomplished. Therefore, this study is quite innovative and not comparable to previous work. Commonly, in the proposed neural network model, the input data is 12 patient features, and the output consists of 5 stages of the liver fibrosis process. However, aiming at our introduced algorithm, the number of features is reduced to 7 inputs with preserved accuracy without significant change. The proposed algorithm reduces the volume of the problem and cost required and optimizes the proposed pattern. In addition to the primary approach of this study, which is the integration of the Neural Network and the TLBO’s innovative algorithm, the dataset has been implemented with Bayesian algorithms, and a decision tree. However, considering the accuracy of liver fibrosis staging in comparison to the previous studies discussed, it is reasonable to make a fair comparison between the results of our current study and the most recent prior research, which achieved the highest diagnostic accuracy on this dataset.

This study deals with the new integration of neural networks and innovative algorithms. The TLBO’s innovative algorithm has been applied in our study. The integration is done in the pre-processing phase and with the help of the TLBO algorithm, feature selection has evolved. This study aimed to introduce a method by which we can predict the possibility of liver fibrosis with fewer blood tests and demographical data. Thus, we proposed a method using a composition of demographical data and blood tests, by which blood donors and patients infected with HCV can be staged for liver fibrosis without invasive procedures, which can reduce the risks and costs. Firstly, owing to the imbalance between healthy and infected cases, we implemented the SMOTE technique. Following this, we used different methods among which Decision tree and deep learning with 12 features produced the highest accuracy, at 0.96. Moreover, the integration of TLBO and MLP methods, with a reduced set of seven primary features, yielded an accuracy rate of 0.89, while the MLP using all 12 features achieved a slightly higher accuracy of 0.903. Considering the preservation of level of accuracy, the composition of this mentioned method with fewereatures is specifically novel in our study compared with previous ones.

In this study, we worked on the dataset which was used by some similar studies previously. An overview about the performance of models used in previous papers have been provided in Table 4.

**Table 4:**
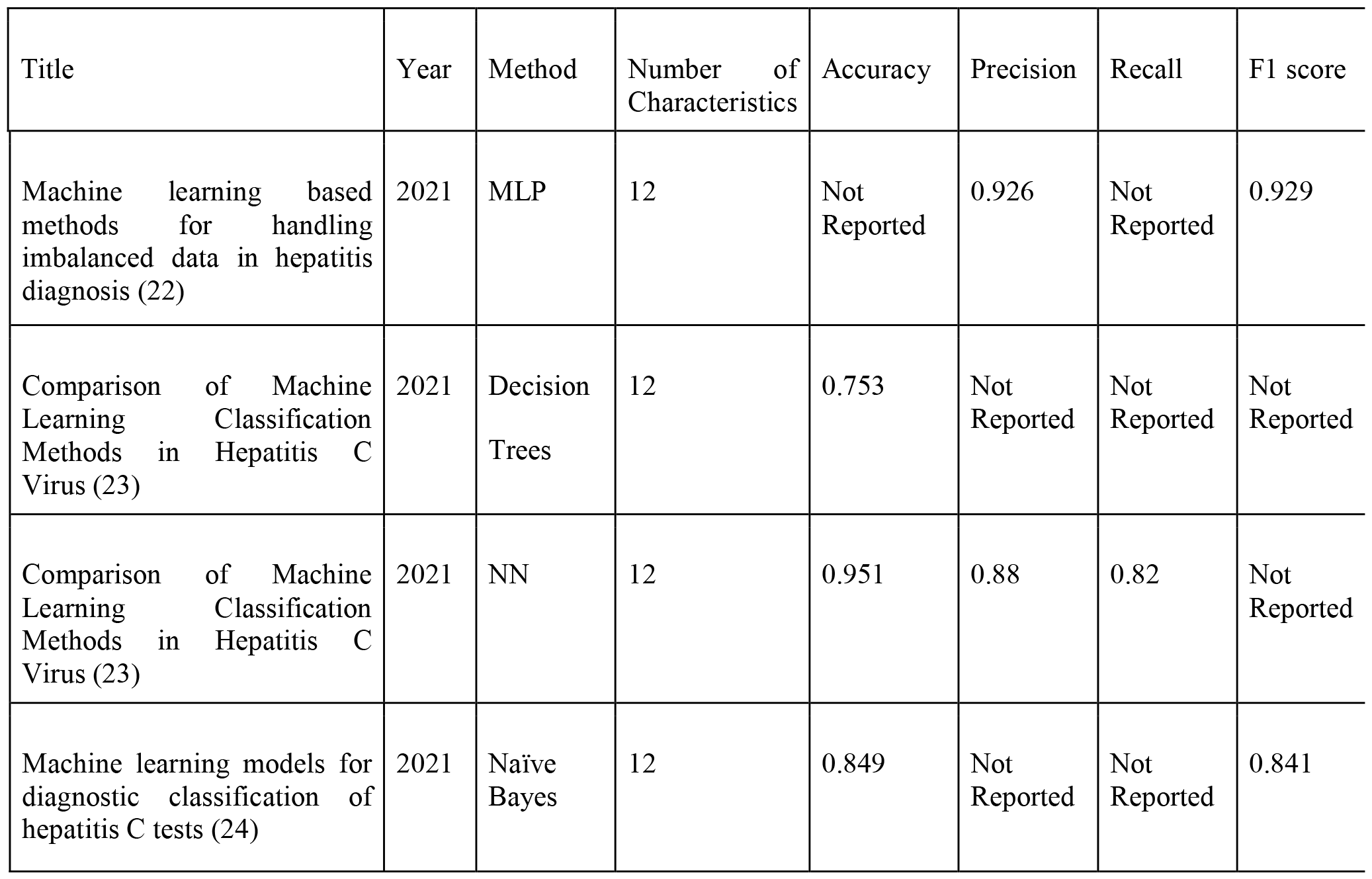
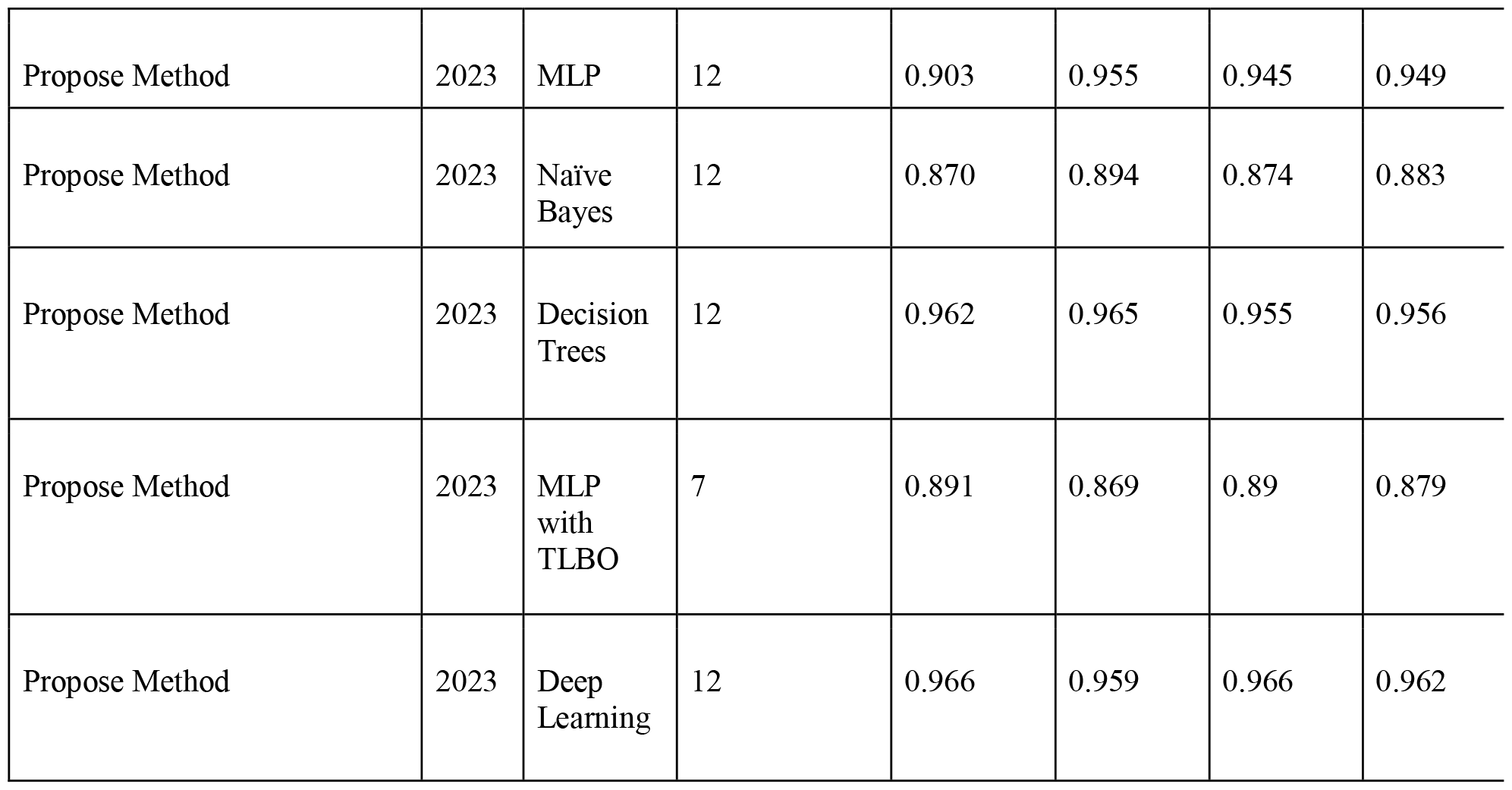
Comparison of the previous studies with our study.

One study was performed using this dataset to recognize an efficient model of machine learning classification for diagnosing liver hepatitis C using 12 features. This study showed that the implementation of SMOTE technique raised performance. In comparison with our study, the integration of SMOTE with NB, KNN, and logistic regression models yielded accuracy rates of 0.84, 0.88, and 0.93 (24). In comparison, our study used the integration of the SMOTE with NB considering 12 features and reached an accuracy rate of 0.855.

Another study on this dataset was carried out using all 12 features by implementing the methods of random forest, MLP, and J48. In the mentioned study, 10-fold cross-validation method was implemented as a method for the evaluation of algorithms. Also, over-sampling and under-sampling methods were used to make a balance in the dataset. Finally, the accuracy levels ranged from 0.65 to 0.99. With respect to MLP method with 12 features, which is a mutual method between our study and the mentioned one, both studies produced about 0.95 of accuracy rate (22).

In 2021, one study used the same dataset but with using 10 features, by performing K-nearest neighbor (KNN), NB, neural network, and random forest. The highest accuracy rate belonged to the neural network at 0.95, which is similar to our result (23). Other methods reached accuracy rates between 0.89 to 0.94, which were not much different from our method.

Considering TLBO implementation, our proposed method is not only more accurate but also can be considered more cost-effective. Our proposed method is able to predict the liver fibrosis stage in blood donors and hepatitis C patients by measuring only seven clinical and demographic features instead of ultrasound guided liver biopsy. Considering the costs and tests’ risks, this method is safe, effective, economical, and time-saving for patients and hospitals compared with the conventional method of liver fibrosis stage detection (25). As a result, fewer blood tests are more likely to be accepted by patients and insurance companies with regard to covering the costs.

### Limitations and future direction

While our research does offer insights into hospital risks and costs, it is imperative to acknowledge our study’s limitations. Considering a limited number of individuals in this study; an increased number of population size and various races and nationalities can affect the results in future studies. Moreover, 12 features are relatively a high number compared with 615 records; we need algorithms with lower numbers of features in future studies. Also, using various patients’ features in the prediction of other HCV outcomes, such as mortality, is highly recommended for future investigations. Also, we were unable to obtain information on liver biopsy for all patients, the specifics of the histology scoring methodology and the platelet counts for calculating FIB-4 index despite our efforts and lack of response from the dataset authors and relevant data on the source website. Finally, the lack of an acceptable external dataset prevents us from performing external validation in this study.

## Conclusion

The methods of Decision trees and deep learning showed the highest levels of accuracy with 12 features. Interestingly, with the use of TLBO and 7 features, the MLP reached a 0.891 accuracy rate which is quite satisfying compared with similar studies. The results between this study and previous studies were examined and showed that the recruited algorithm of our study was more straightforward, with lower required properties and higher accuracy, which can lead to having a diagnostic method with minimal cost and lower required patient information.

## Supporting information

Title Page

## Data Availability

All data produced in the present study are available upon reasonable request to the authors

## Acknowledgment

None

