## Supplementary material for "An Integrated Neural Network and Evolutionary Algorithm Approach for Liver Fibrosis Staging: Can Artificial Intelligence Reduce Patient Costs?": Title Page

**Running title**: Artificial Intelligence for Liver Fibrosis Staging

**Authors’ names and affiliations:**

Ali Nazarizadeh^1^, Touraj Banirostam^1^, Taraneh Biglari^1^, Mohammadreza Kalantarhormozi^2^, Fatemeh Chichagi^3^, Amir Hossein Behnoush^4^, Mohammad Amin Habibi^5^, Ramin Shahidi^6^

1-      Department of Computer Engineering, Central Tehran Branch, Islamic Azad University, Tehran, Iran

a.
b.
c.

2-      The Persian Gulf Tropical Medicine Research Center, The Persian Gulf Biomedical Sciences Research Institute, Bushehr University of Medical Sciences, Bushehr, Iran

a.

3-      Students' Scientific Research Center (SSRC), Tehran University of Medical Sciences, Tehran, Iran

a.

4-      Non–Communicable Diseases Research Center, Endocrinology and Metabolism Population Sciences Institute, Tehran University of Medical Sciences, Tehran, Iran

a.

5-      Clinical Research Development Center, Shahid Beheshti Hospital, Qom University of Medical Sciences, Qom, Iran

a.

6-      School of Medicine, Bushehr University of Medical Sciences, Bushehr, Iran

a.

**Corresponding author's contact information:**

Ramin Shahidi, MD.

School of Medicine, Bushehr University of Medical Sciences, Bushehr, Iran

Address: Bushehr University of Medical Sciences, Moallem St, Bushehr, Iran.

Tell: +98-77-33321621

ORCID ID: 0000-0003-1039-1407

**Acknowledgment:** None

**Data availability statement:** The data is available within the article or is achievable through the request of the corresponding author.

**Funding statement:** The present study has not received any funding.

**Conflict of interest disclosure:** The authors declared that no conflict of interest existed.

**Ethics approval statement:** This article contains no patient-identifying information, so ethical approval is not required.
